# Next-generation DNA sequencing identifies mutated FANCD2 Gene as a novel Biomarker for monitoring early disease progression and timely therapeutic interventions in advanced phase Chronic Myeloid Leukemia patients

**DOI:** 10.1101/2023.12.19.23300103

**Authors:** Nawaf Alanazi, Abdulaziz Siyal, Muhammad Absar, Masood Shammas, Sarah Al-Mukhaylid, Amer Mahmood, Zafar Iqbal

**Author notes:** **Corresponding authors: Dr. Amer Mahmood and Dr. Zafar Iqbal,**.

## Abstract

Chronic Myeloid Leukemia, resulting due to chromosomal aberration t(9;22) through formation of oncogenic BCR-ABL fusion oncogene. Modern BCR-ABL inhibitors, called TKIs, have revolutionized CML treatment. CML has three stages: chronic, accelerated, and blast crisis. TKIs work well in CP-CML, where patients survive as long as the normal population, but they don’t work in AP- and BC-CML. Even with advances in treatment, BC-CML has an average overall survival of less than a year, giving oncologists little time to clinically intervene. Oncologists can delay or prevent CML advancement by detecting patients at risk of disease progression early and making timely treatment decisions, especially with third and fourth generation TKIs. However, no universal molecular biomarkers exist to diagnose CML patient groups at risk of disease progression.

A recent study found that all BC-CML patients have mutant FANCD2. Our study was designed to detect mutant FANCD2 in AP-CML (early progression phase) to investigate its potential as a novel biomarker of early CML progression from chronic phase to accelerated phase due to the urgent need for such a biomarker.

Our study comprised of 123 CP-CML (control group) and 60 AP-CML patients (as experimental group) from Hayatabad Medical Complex, Peshawar, Pakistan, from Jan 2020 to July 2023. DNA was extracted from the patients and FANCD2 gene was sequenced using Illumina next generation sequencer (NGS) Illumina MiSeq sequencer. NGS analysis revealed a unique splice site mutation in FANCD2 gene (c. 2022-5C>T). This mutation was detected in all CP-CML patients but in none of CP-CML. The mutation was confirmed by Sanger sequencing.

FANCD2 is member of Fanconi anemia (FA-) pathway gene involved in DNA repair and genomic instability. Therefore, our studies show that FANCD2 (c. 2022-5C>T) mutation as a very specific molecular biomarker for early CML progression. We recommend to clinical validate this biomarker is prospective clinical trials.

## 1. Introduction

Chronic Myeloid Leukemia (CML), is a chronic myeloproliferative malignancy of stem cells that is manifested in the blood (Senapati et al. 2023). It is caused by t(9;22), leading to formation of a chromosomal abnormality called as Philadelphia (Ph) chromosome (Narlı et al. 2023). This chromosomal abnormality results in BCR-ABL fusion oncogene responsible for onset of cancer in myeloid lineage of hematopoietic stem cells (Eden et al. 2023). Estimates of the annual prevalence range from 0.6 to 2.0 instances per 100,000 people monitored, or around 10-15% of newly diagnosed adult cases of leukemia (Sasaki et al, 2023, Siegel et al, 2017). The median age of diagnosis for CML is between 57 and 60 years old, and 1.2-1.7% more males than women get the disease (Apperley, 2015; Jabbour, 2020, Cortes, 2006, Lurlo et al, 2023).

The modern BCR-ABL inhibitor drugs called tyrosine kinase inhibitors (TKIs) CML have revolutionized CML treatment. However, there are certain challenges in managing patients with these drugs (Senapati et al. 2023). CML has three disease phases called as chronic phase (CP-), accelerated phase (AP-) and blast crisis (BC-) CML (Boucher et al., 2023). TKIs are very effective in CP-CML, resulting in the overall survival rate improving from 20% to 80–90% (Venkitaraman, 2004; Senapati et al. 2023). This led to overall survival of CP-CML patients equal to general public, at least in technologically advanced countries like USA, Canada, Europe and Japan (Busch et al, 2023). Nevertheless, patients in early progression phase (AP-CML) and terminal progression phase (BC-CML) show resistance to TKIs (Takahashi, 2023). Specifically in BC-CML, despite all advancements in treatment modalities, the average overall survival is less than a year, which provides a little time frame for oncologists to clinically intervene CML progression (Yoshimaru & Minami,, 2023). Early detection of CML patients at risk of disease progression can considerably help oncologists to delay or even avoid CML progression by timely treatment decision making specifically with the introduction of third and fourth generation TKIs (Shin et al., 2023). Nevertheless, no specific and universal molecular biomarkers exist for timely detection of CML patient groups at risk of disease progression (Li et al. 2023).

Cancer has several mechanisms involved in the initiation and progression of its different types, including DNA repair defects leading to genomic instability (Telliam et al., 2023). Fanconi anemia (FA) is a rare autosomal recessive hereditary illness characterized by gene mutations that are predominantly involved in DNA damage response or repair (Leung et al., 2023). The FANC genes play a crucial role in the FA pathway, regulating DNA damage responses through complicated processes like ubiquitination, phosphorylation, and degradation signals, all of which are required for genome stability and genomic integrity (Dong, 2015). Due to increased genomic instability, it is well known that the FANC gene dysfunction increases the chance of developing various hematological and solid malignancies (D’Andrea, 2003). FANCD2 mutations have recently been found associated with BC-CML as a biomarker of terminal CML progression (Absar et al, 2020). Accordingly, this study was designed to find out the potential of mutated FANCD2 as a biomarker of early CML progression in AP-CML. For the first time in literature, we hereby report mutations in major FA-pathway genes associated exclusively with early CML progression in AP-CML patients.

## 2. Materials and Methods

### Patient selection and recruitment

Clinical Follow-up The study was conducted on CML patients enrolled in Hayatabad Medical Complex (HMC) Peshawar, Khyber Pakhtunkhawa (KP), Pakistan, from January 2020 to July 2023. The number of CP-CML patients included in this study was 123. The experimental group compromised 60 Accelerated Phase (AP-CML) patients, while 123 age/gender-matched Chronic Phase (CP-CML) patients were controls. All patients were initially treated with imatinib mesylate (IM), and patients with IM resistance were given nilotinib (NI). The European Leukemia Net guidelines 2020 were utilized to determine the criteria of treatment responses (Baccarani, 2013; Baccarani, 2015, Yilmaz et. 2023). Standard terminologies, version 4.03 was used to classify the hematological adverse events and other adverse events (Cortes, 2012).

The regulations of the Declaration of Helsinki were followed throughout the study. All patients included in the study provided written informed consent (World Medical Association, 2007; Goodyear, 2007). The approval of study protocols was obtained from Scientific Committees and Ethical Review Boards (ERBs) of King Abdullah International Medical Research Center (KAIMRC); King Saud bin Abdulaziz University for Health Sciences (KSAU-HS), Hayatabad Medical Complex (HMC), Peshawar, Pakistan; and University of the Punjab, Lahore, Pakistan.

### Sample Collection and DNA Extraction

Peripheral blood samples were collected in 3-5ml EDTA tubes (BD Vacutainer Systems, Franklin Lakes, N.J.) from all age groups and clinical phases of CML patients and stored at - 70°C for further examination. Venous blood samples were obtained from registered CML patients biweekly, for follow-up and medication refills, during their visits to the outpatient department (OPD) of the medical oncology unit, HMC, Peshawar Khyber Pakhtunkhawa (KPK) Pakistan. All blood samples, DNA extraction kits, and reagents were set to room temperature (15-25°C) prior to DNA extraction by using a 56°C water bath. The extraction of genomic DNA from blood samples was performed using the QIAamp DNA Mini Kit (#51306) (Qiagen).

### Targeted Resequencing of FANCD2 using Next Generation Sequencing (NGS)

To represent each clinical phase of the disease (Chronic Phase and Accelerated Phase), well-characterized CML samples were selected and processed for NGS (Gnirke, 2009). Illumina® DNA Prep with Enrichment, (S) Augmentation kit (Cat. # 20025523) was utilized for target enrichment (Gnirke, 2009; AlAsiri, 2015). The first step to NGS was DNA fragmentation, followed by tagmentation. Afterward, tagmented DNA fragments were amplified and then purified using magnetic beads. Next, Oligos were utilized to capture target regions. Enriched libraries were amplified by PCR and quantified using a Qubit fluorometer, while Agilent Bioanalyzer was equipped to measure the library size distribution. Finally, cluster generation and exon sequencing were performed using the Illumina NextSeq500 instrument by loading the quantified DNA libraries on the flow cell (AlAsiri, 2015, Absar et al., 2020).

### Next Generation Sequencing (NGS) Data Analysis

The conversion of output files, BCL files to FASTQ files was done by BCL2FASTQ software. Alignment of FASTQ files to the human genome was performed by BWA Aligner, applying the BWA-MEM algorithm. Variants were called by the Genome analysis tool kit (GATK). Illumina Variant Studio was used for the annotation and filtration of genomic variants (AlAsiri, 2015).

### Primary Analysis

FANCD2 gene was analyzed in all AP-CML patients to detect shared biomarkers of CML progression. Filtration strategies that relied on calling rare variants and excluding intron and synonymous variants were applied to modify the excel file presenting NGS. Furthermore, all variants with known prediction were removed, either benign (B) or tolerant (T). Some variants were considered as B when it has 70% or more of B, while other variants were classified as T when T’s frequency was 70% or more (Carson, 2014). Variants with more than 0.005 population frequency in the dbSNP and Exome Sequencing Project (ESP) database were also eliminated. Thus, variant calling was only limited to variants with intermediate and high protein effects along with splice variants, resulting in about 124 rare variants. Moreover, data was further analyzed to investigate novel gene mutations that are present in AP-CML patients but not in CP-CML patients and healthy controls, suggesting its role in disease progression (Branford, 2018; Xu, 2020). Access to data made by next-generation sequencing can be obtained from NCBI, to which it was submitted, at https://www.ncbi.nlm.nih.gov/sra/PRJNA734 750 (SRA accession number PRJNA734750; accessed on 28 September 2022).

### Validation of Mutation by Sanger Sequencing

Samples were prepared using ABI Prism 3730 Genetic Analyzer (Applied Biosystems, California, USA) and ABI PRISM Big Dye Terminator Cycle Sequencing Ready Reaction kits, and Amplification of samples was done via PCR. Variants identified through NGS were validated by Sanger sequencing (Tsiatis, 2010). Forward and reverse sequencing of PCR amplified FANCD2 fragments were performed by the Sanger sequencer and mutational analysis carried out, as described earlier (Absar et al. 2020).

### Statistical Analysis of Patient Clinical Data

Categorical variables were represented with percentages and absolute numbers, while continuous variables were measured with mean and median according to the normality test. Chi-Square and Fisher’s exact test were utilized to compare categorical data of two groups, depending on Applicability, while the comparison of two groups of continuous data was done by Two sample independent test or Mann Whitney U test, depending on the normality hypothesis. ANOVA or Kruskal-Wallis test were performed to analyze data from more than 3 groups. Assessment of survival outcome was made using Kaplan-Meier survival analysis curves, and the log-rank test was used to compare groups. [SAS/STAT] software version 9.4 (SAS Institute Inc., Cary, NC, USA.) and R foundation were used for data analysis and statistical computing (Vienna, Austria), accordingly (R Core Team., 2012). Calculations of the Sokal risk score, Eutos risk score, and Euro risk scores were also done (Sokal, 1984; Hasford, 1998; Hasford, 2011).

## 3. Results

This study comprised 183 CML patients. The overall mean age of all patients was 34.6. However, mean age for CP-CML and AP-CML patients was 33.5 and 35.6, respectively. Regarding gender, CML was more common in males as they constituted 60.5%, while females were only 39.5%, giving a significant male-to-female ratio of 1.6:1(p = 0.0200). Moreover, the male-to-female ratios for CP-CML and AP-CML patients were 1.5:1 and 2:1, accordingly. The means of clinical characteristics calculated were 10.1 for hemoglobin, 317.9 for white blood cell count, and 400.2 for platelet count. Furthermore, anemia and leukocytosis of more than 50 × 109/L were observed in more than two third of the patients. In addition, various types of treatment were applied to patients, including Imatinib and chemotherapy. Overall, characteristics including male-to-female ratio, hemoglobin level, WBC count, platelet count, treatment type, hepatomegaly, splenomegaly, and survival status were significantly altered in AP-CML patients, compared to CP-CML patients. The comparison between CML phases in regard to patients’ demographic and laboratory characteristics are displayed in table 1.

**Table 1:**
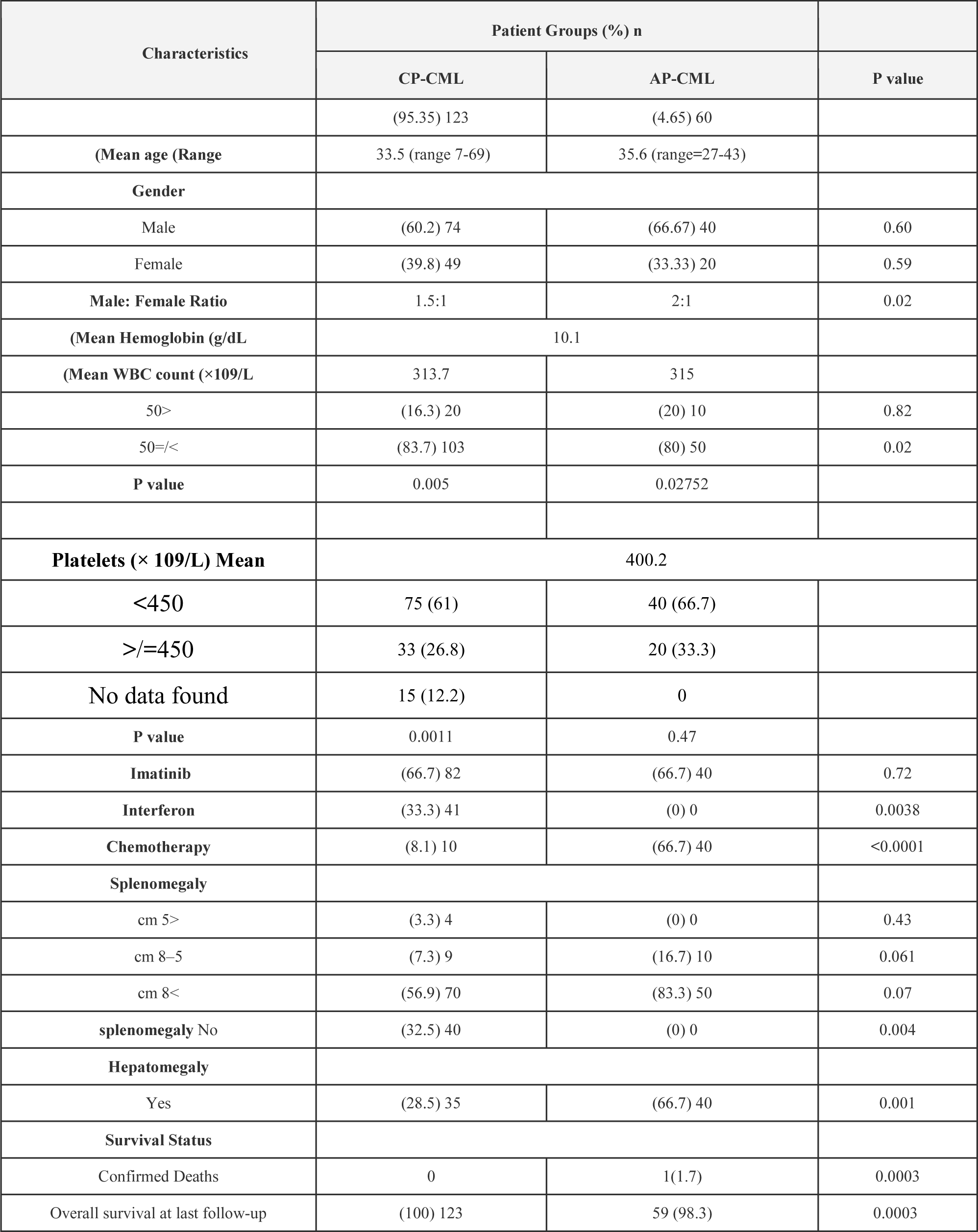
A comparison between CP- and AP-CML patients in this study in regard to their demographic and laboratory characteristics.

### Next Generation Sequencing (NGS)

The results of NGS indicated a novel splice site mutation at genomic position 10,106,408, corresponding to leading to cytosine to thymine substitution (c. 2022-5C>T), in the FANCD2 gene. This gene is an important member of FA-pathway genes located in chromosome 3. This mutation was shared by all AP-CML patients but not CP-CML patients, suggesting its association with disease progression.

### Validation by Sanger Sequencing

A heterozygous variant (C2022T) was found and validated by sanger sequencing. The FANCD2, c. 2022-5C>T (genomic position 10,106,408) detected by NGS was confirmed by Sanger sequencing as indicated in Figure 3. This demonstrates that the above-mentioned mutation is an indicator of early CML progression in CML patients.

As a result of our clinical validation investigations, it is concluded that the FANCD2 gene is exclusively mutated in all AP-CML patients but not in any of the control subjects. Our research indicates that the FANCD2 (c. 2022-5C>T) mutation can serve as a highly specific molecular biomarker for the early progression of CML.

## 4. Discussion

In our study, 4.65% of 129 CML patients progressed to AP-CML. We found that the FANCD2 gene was mutated in all patients in the accelerated phase, compared to healthy individuals and Chronic phase CML patients. To consolidate our findings, we found another study suggesting that the crucial FA pathway protein, FANCD2, has been linked to the advancement of the CML illness (Valeri, 2012). In 2007, a study concluded that the frequency of FANCD2 was greater than that of any other FA complementation group (Kalb, 2007). Although FANCD2’s exact function is yet unclear, the upkeep of genomic stability is assumed to be mostly regulated by FANCD2(31). According to Valeri et al., BCR-ABL1 induces genetic instability in CML cells by inhibiting FANCD2 nuclear foci, leading to centrosomal amplification and DNA repair deficiencies. FANCD2, located at 3p25.3, plays a role in repairing impaired DNA (Valeri, 2012; Meetei, 2003). However, a mutated FANCD2 gene will result in its inability to continue the DNA-repairing cascade (Absar, 2023). The FANCD2 variant detected in this study was located in chromosome 3, and it was positioned at 10106408 along with a displacement of Cytosine to Thymine. By the end of 2015, A similar FANCD2 variant, NM_001018115.3(FANCD2): c.2022-5C>T), had its first publication. Conflicting theories about the pathogenicity of both comparable variants were present. They have been associated with FA as a pathogenic and with Hereditary Breast Ovarian Cancer Syndrome as benign (National Center for Biotechnology Information).

In our study, the splice site mutation between intron 22 and exon 23 resulted in the intron 22 variant. The function of the intron 22 variant on FANCD2 mono-ubiquitination remains uncertain, but it could be predicted since it is close to intron 19, which is the FANCD2 mono-ubiquitination site (Lewis, 2005). A study in China identified a missense mutation c.3713T>A; p.M1238K in the FANCD2 gene that leads to non-expression of the FANCD2 protein. Furthermore, function studies were applied to find that other splice site mutations in the FA gene cause exon skipping (Li, 2018). Another study in the US detected 25 intronic variants and 6 silent coding variants that lead to familial breast cancer. One of which was in exon 23 c.2148 C>G, resulting in T716T protein change (Lewis, 2005).

The FANCI-FANCD2 heterodimer is where the FA pathway comes together (Wang, 2004). It serves as a substrate for the FA core complex as well as a potential collecting site for proteins involved in downstream DNA repairs, such as FAN1 nuclease and other FANC proteins (MacKay, 2012). Despite only possessing a 14% conservation in their solenoidal structures, FANCI and FANCD2 were known to have striking similarities in the 2011 discovery of the crystal lattice of mouse FANCI-FANCD2 (Joo, 2011). More than 97% of FA patients have a deficiency caused by mutations in the genes encoding FANCD2 and FANCI (Wang, 2015). At the region of DNA damage, the FA proteins function as a ubiquitin E3 ligase to monoubiquitinate the FANCI-FANCD2 pair (van Twest, 2017). This results in enlisting downstream nucleases with ubiquitin-binding domains to restore the interstrand DNA bridge (Smogorzewska, 2010). Although monoubiquitination and FA pathway activation necessitate DNA binding of FANCI-FANCD2 (Liang, 2016). It is not certain how it triggers the repair of DNA (Li, 2020).

High FA gene expression is typically associated with chemo-resistance; the high expression level of FANCD2 is associated with reduced chemotherapy sensitivity and a higher tumor mutation rate. It was observed in breast, lung, and ovarian cancers, Thus, resulting in reduced survival time (Dan, 2021; Miao, 2022). FA pathway inhibition by targeted therapies is a promising approach for improving the efficacy of chemotherapy due to its role in chemoresistance across a wide range of cancers (Liu et al., 2020). In early studies, Curcumin, Wortmannin, H-9, and Alsterpaullone were found to inhibit FANCD2 by apoptosis through the NFκB pathway (Chirnomas, 2006). A study further assessed monoketone analogues of Curcumin and found that EF24 was more specific and active against monoubiquitination of FANCD2 (Shen et al. 2015). Lastly, a recent study has identified CU2 as a compound that shows potential biochemical ubiquitylation selectivity and activity against the FA pathway (Cornwell, 2019). Currently, three PARP inhibitors that target the FA pathway are FDA-approved for treating relapsed breast and ovarian cancers, olaparib, rucaparib, and niraparib (Niraj, 2019).

In addition, there have been reports that linked cancer incidence with FA pathway mutations (Shen, 2015). The previously mentioned mutations were reported to cause FA bone marrow failure. Also, the most well-known genes that predispose to breast cancer are BRCA1 and BRCA2, both of which are considered part of the FANC gene family. This system is frequently referred to as the FA-BRCA pathway given the growing relationship between FA and the genes for breast cancer (Williams, 2011). In addition, two studies were investigating the relationship between FANCD2 and breast cancer. In China, a poor prognosis was observed in sporadic breast cancer patients with high levels of FANCD2 (Feng, 2019). A study in Finland supports these findings by showing a significant association of variant (c.2715 + 1G > A) in the FANCD2 gene with breast cancer (Mantere, 2017). Also, studies conducted in the United Kingdom and Poland on ovarian carcinoma samples verified that the risk of recurrence and death are highly associated with the expression level of FANCD2 (Moes-Sosnowska, 2019; Mani, 2021). On the other hand, in the United States, a study conducted on 181 ovarian cancer patients provided evidence of an increased survival rate in patients with FANCD2 mutation (Joshi, 2020). On the basis of its upregulation, FANCD2 has an unsatisfactory prognosis for other types of cancer, which may contribute to tumorigenesis. Such cancers are esophageal squamous cell carcinoma, head and neck carcinoma, nasopharyngeal carcinoma, and lung adenocarcinoma (Lei, 2020; Chandrasekharappa, 2017; Xu, 2019).

In addition, the FANCD2 gene mutation has also been observed in non-cancer diseases. Two studies have proven the association of FANCD2 mutation with other diseases. The first one was a case report of a 2.5-year-old Saudi boy from Al-Ahsa diagnosed with ambiguous genitalia and intrauterine growth restriction (IUGR). This study found that ambiguous genitalia in Saudi male infants could be related to c.2605 + 1G>A variant resulting from a homozygous mutation in the FANCD2 gene. The second study revealed that the FANCD2 expression level is significantly increased in Glioblastoma patients. Moreover, it also plays a role in drug resistance and the progression of the disease (Xu et al., 2023). The impact of FANCD2 mutation on CML progression and drug resistance was evident as discussed above. This highlights the significance of understanding the relationship between FANCD2 and CML (Leung et al., 2023; Valeri net al., 2023).

## 5. Conclusions

We discovered that the FANCD2 gene was mutated in every AP-CML patient. FANCD2 is member of Fanconi anemia (FA-) pathway gene involved in DNA repair and genomic instability. Therefore, our studies show that FANCD2 (c. 2022-5C>T) mutation as a very specific molecular biomarker for early CML progression. We recommend to clinical validate this biomarker is prospective clinical trials.

**Figure 1:**
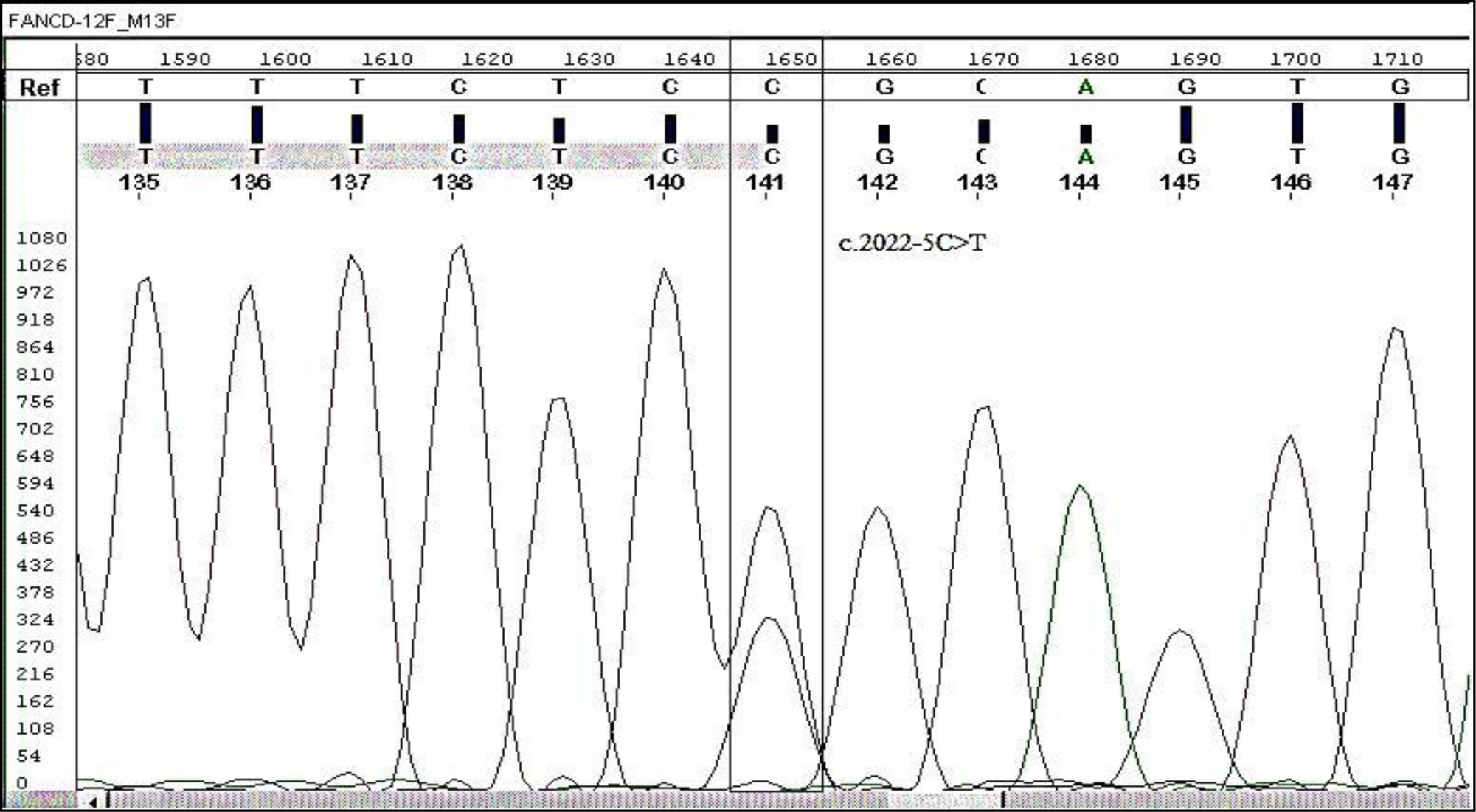
Electropherogram showing mutation C > T in FANCD2 at splice site of intron 22 and its comparison with reference / wild-type sequence

## Data Availability

All data produced in the present study are available upon reasonable request to the authors

## 7. Acknowledgements

The study was approved by King Abdullah International Medical Research Centre (KAIMRC), National Guard Health Affairs, Saudi Arabia, although no research funding was provided (project # RA17/002/A). This study was partially supported by the College of Medicine, Research Centre, Deanship of Scientific Research, King Saud University, Riyadh, Saudi Arabia. All authors have read and have consented to the acknowledgement.

## Research Ethics’ Statements and Approval of the study

The regulations of the Declaration of Helsinki were followed throughout the study. All patients included in the study provided written informed consent (World Medical Association, 2007; Goodyear, 2007).

The approval of study protocols was obtained from Scientific Committees and Ethical Review Boards (ERBs) of King Abdullah International Medical Research Center (KAIMRC); King Saud bin Abdulaziz University for Health Sciences (KSAU-HS), Hayatabad Medical Complex (HMC), Peshawar, Pakistan; and University of the Punjab, Lahore, Pakistan.

## References

1. D’Andrea, A. D., & Grompe, M. (2003). The Fanconi anaemia/BRCA pathway. *Nature Reviews Cancer*, 3(1), 23–34. doi: 10.1038/nrc970.

2. Absar, M., Mahmood, A., Akhtar, T., Basit, S., Ramzan, K., Jameel, A., … & Iqbal, Z. (2020). Whole exome sequencing identifies a novel FANCD2 gene splice site mutation associated with disease progression in chronic myeloid leukemia: Implication in targeted therapy of advanced phase CML. *Pakistan Journal of Pharmaceutical Sciences*, 33(3(Special)), 1419–1426. PMID: 33361032.

3. AlAsiri, S., Basit, S., Wood-Trageser, M., Yatsenko, S. A., Jeffries, E. P., Surti, U., … & Haque, M. F.-U. (2015). Exome sequencing reveals MCM8 mutation underlies ovarian failure and chromosomal instability. *Journal of Clinical Investigation*, 125(1), 258–262.

4. Ali, M. A. (2016). Chronic Myeloid Leukemia in the Era of Tyrosine Kinase Inhibitors: An Evolving Paradigm of Molecularly Targeted Therapy. Molecular Diagnosis & Therapy, 20(4), 315–333. doi: 10.1007/s40291-016-0208-1.

5. Apperley, J. F. (2015). Chronic myeloid leukaemia. The Lancet, 385(9976), 1447–1459. doi: 10.1016/S0140-6736(13)62120-0.

6. Baccarani, M., Castagnetti, F., Gugliotta, G., & Rosti, G. (2015). A review of the European LeukemiaNet recommendations for the management of CML. *Annals of Hematology*, 94(Suppl 2), S141–S147.

7. Baccarani, M., Deininger, M. W., Rosti, G., Hochhaus, A., Soverini, S., Apperley, J. F., … & Hehlmann, R. (2013). European LeukemiaNet recommendations for the management of chronic myeloid leukemia: 2013. *Blood*, 122(6), 872–884.

8. Branford, S., Wang, P., Yeung, D. T., Thomson, D., Purins, A., Wadham, C., … & Hughes, T. P. (2018). Integrative genomic analysis reveals cancer-associated mutations at diagnosis of CML in patients with high-risk disease. *Blood*, 132, 948–961.

9. Carson, A. R., Smith, E. N., Matsui, H., Brækkan, S. K., Jepsen, K., Hansen, J.-B., … & Frazer, A. K. (2014). Effective filtering strategies to improve data quality from population-based whole exome sequencing studies. *BMC Bioinformatics*, 15, 125.

10. Chandrasekharappa SC, Chinn SB, Donovan FX, Chowdhury NI, Kamat A, Adeyemo AA, Thomas JW, Vemulapalli M, Hussey CS, Reid HH, Mullikin JC, Wei Q, Sturgis EM. Assessing the spectrum of germline variation in Fanconi anemia genes among patients with head and neck carcinoma before age 50. Cancer. 2017 Oct 15;123(20):3943–3954. doi: 10.1002/cncr.30802. Epub 2017 Jul 5. PMID: 28678401; PMCID: PMC5853120.

11. Chirnomas D, Taniguchi T, de la Vega M, et al. Chemosensitization to cisplatin by inhibitors of the Fanconi anemia/BRCA pathway. Mol Cancer Ther. 2006;5(4):952–961. doi:10.1158/1535-7163.MCT-05-0493

12. Chronic granulocytic leukaemia: comparison of radiotherapy and busulphan therapy. Report of the Medical Research Council’s working party for therapeutic trials in leukaemia. (1968). British Medical Journal, 1(5586), 201–208. doi: 10.1136/bmj.1.5586.201.

13. Cornwell, M. J., Thomson, G. J., Coates, J., Belotserkovskaya, R., Waddell, I. D., Jackson, S. P., & Galanty, Y. (2019). Small-Molecule Inhibition of UBE2T/FANCL-Mediated Ubiquitylation in the Fanconi Anemia Pathway. ACS chemical biology, 14(10), 2148–2154. 10.1021/acschembio.9b00570

14. Cortes, E. J., Lipton, J. H., Miller, C. B., Ailawadhi, S., Akard, L., Pinilla-Ibarz, J., … & Mauro, M. J. (2012). Change in Chronic Low-Grade Nonhematologic Adverse Events (AEs) and Quality of Life (QoL) in Adult Patients (pts) with Philadelphia Chromosome–Positive (Ph+) Chronic Myeloid Leukemia in Chronic Phase (CML-CP) Switched From Imatinib (IM) to Nilotinib (NIL). *Blood*, 120(21), 3782.

15. Cortes, J. E., Talpaz, M., O’Brien, S., Faderl, S., Garcia-Manero, G., Ferrajoli, A., … Kantarjian, H. M. (2006). Staging of chronic myeloid leukemia in the imatinib era: an evaluation of the World Health Organization proposal. Cancer, 106(6), 1306–1315. doi: 10.1002/cncr.21756.

16. Dan, C., Pei, H., Zhang, B. et al. Fanconi anemia pathway and its relationship with cancer. GENOME INSTAB. DIS. 2, 175–183 (2021). 10.1007/s42764-021-00043-0

17. Dong, H., Nebert, D. W., Bruford, E. A., Thompson, D. C., Joenje, H., & Vasiliou, V. (2015). Update of the human and mouse Fanconi anemia genes. Human Genomics, 9, 32. doi: 10.1186/s40246-015-0054-y.

18. Feng L, Jin F. Expression and prognostic significance of Fanconi anemia group D2 protein and breast cancer type 1 susceptibility protein in familial and sporadic breast cancer. Oncol Lett. 2019 Apr;17(4):3687–3700. doi: 10.3892/ol.2019.10046. Epub 2019 Feb 18. PMID: 30881493; PMCID: PMC6403512.

19. Gnirke, A., Melnikov, A., Maguire, J., Rogov, P., LeProust, E. M., Brockman, W., … & Nusbaum, C. (2009). Solution hybrid selection with ultra-long oligonucleotides for massively parallel targeted sequencing. *Nature Biotechnology*, 27(2), 182–189. doi: 10.1038/nbt.1523.

20. Goodyear, E., Krleza-Jeric, M. D., & Lemmens, K. (2007). The Declaration of Helsinki. *BMJ*, 335, 624–625.

21. Hasford, J., Baccarani, M., Hoffmann, V., Guilhot, J., Saussele, S., Rosti, G., … & Hehlmann, R. (2011). Predicting complete cytogenetic response and subsequent progression-free survival in 2060 patients with CML on imatinib treatment: the EUTOS score. *Blood*, 118(3), 686–692.

22. Hasford, J., Pfirrmann, M., Hehlmann, R., Allan, N. C., Baccarani, M., Kluin-Nelemans, J. C., … & Ansari, H. (1998). Writing Committee for the Collaborative CML Prognostic Factors Project Group. A new prognostic score for survival of patients with chronic myeloid leukemia treated with interferon alfa. *Journal of the National Cancer Institute*, 90(11), 850–858.

23. Jabbour, E., & Kantarjian, H. (2020). Chronic myeloid leukemia: 2020 update on diagnosis, therapy and monitoring. American Journal of Hematology, 95(6), 691–709. doi: 10.1002/ajh.25792.

24. Joo W, Xu G, Persky NS, Smogorzewska A, Rudge DG, Buzovetsky O, … & Pavletich NP. (2011). Structure of the FANCI-FANCD2 complex: insights into the Fanconi anemia DNA repair pathway. *Science*, 333(6040), 312–316. doi: 10.1126/science.1205805.

25. Joshi S, Campbell S, Lim JY, McWeeney S, Krieg A, Bean Y, Pejovic N, Mhawech-Fauceglia P, Pejovic T. Subcellular localization of FANCD2 is associated with survival in ovarian carcinoma. Oncotarget. 2020 Feb 25;11(8):775–783. doi: 10.18632/oncotarget.27437. PMID: 32165999; PMCID: PMC7055545.

26. Kalb R, Neveling K, Hoehn H, Schneider H, Linka Y, Batish SD, Hunt C, Berwick M, Callen E, Surralles J, Casado JA, Bueren J, Dasi A, Soulier J, Gluckman E, Zwaan CM, van Spaendonk R, Pals G, de Winter JP, Joenje H, Grompe M, Auerbach AD, Hanenberg H, Schindler D. (2007). Hypomorphic mutations in the gene encoding a key Fanconi anemia protein, FANCD2, sustain a significant group of FA-D2 patients with severe phenotype. *American Journal of Human Genetics*, 80(5), 895–910. doi: 10.1086/517616.

27. Landais, I., Hiddingh, S., McCarroll, M., Yang, C., Sun, A., Turker, M. S., Snyder, J. P., & Hoatlin, M. E. (2009). Monoketone analogs of curcumin, a new class of Fanconi anemia pathway inhibitors. Molecular cancer, 8, 133. 10.1186/1476-4598-8-133

28. Lei LC, Yu VZ, Ko JMY, Ning L, Lung ML. FANCD2 Confers a Malignant Phenotype in Esophageal Squamous Cell Carcinoma by Regulating Cell Cycle Progression. Cancers (Basel). 2020 Sep 7;12(9):2545. doi: 10.3390/cancers12092545. PMID: 32906798; PMCID: PMC7565464.

29. Lewis AG, Flanagan J, Marsh A, Pupo GM, Mann G, Spurdle AB, Lindeman GJ, Visvader JE, Brown MA, Chenevix-Trench G; Kathleen Cuningham Foundation Consortium for Research into Familial Breast Cancer. (2005). Mutation analysis of FANCD2, BRIP1/BACH1, LMO4 and SFN in familial breast cancer. *Breast Cancer Research*, 7(6), R1005–16. doi: 10.1186/bcr1336.

30. Li L, Tan W, Deans AJ. Structural insight into FANCI-FANCD2 monoubiquitination. Essays Biochem. 2020 Oct 26;64(5):807–817. doi: 10.1042/EBC20200001. PMID: 32725171; PMCID: PMC7588663.

31. Li N, Ding L, Li B, Wang J, D’Andrea AD, Chen J. (2018). Functional analysis of Fanconi anemia mutations in China. *Experimental Hematology*, 66, 32–41.e8. doi: 10.1016/j.exphem.2018.07.003.

32. Liang CC, Li Z, Lopez-Martinez D, Nicholson WV, Vénien-Bryan C, Cohn MA. The FANCD2-FANCI complex is recruited to DNA interstrand crosslinks before monoubiquitination of FANCD2. Nat Commun. 2016 Jul 13;7:12124. doi: 10.1038/ncomms12124. PMID: 27405460; PMCID: PMC4947157

33. Liu W, Palovcak A, Li F, Zafar A, Yuan F, Zhang Y. Fanconi anemia pathway as a prospective target for cancer intervention. Cell Biosci. 2020;10:39. Published 2020 Mar 16. doi:10.1186/s13578-020-00401-7

34. MacKay C, Déclais AC, Lundin C, Agostinho A, Deans AJ, MacArtney TJ, … & Rouse J. (2010). Identification of KIAA1018/FAN1, a DNA repair nuclease recruited to DNA damage by monoubiquitinated FANCD2. *Cell*, 142(1), 65–76. doi: 10.1016/j.cell.2010.06.021.

35. Mahon, F. X., & Etienne, G. (2014). Deep molecular response in chronic myeloid leukemia: the new goal of therapy? *Clinical Cancer Research*, 20(2), 310–322. doi: 10.1158/1078-0432.CCR-13-1988.

36. Mani C, Tripathi K, Chaudhary S, Somasagara RR, Rocconi RP, Crasto C, Reedy M, Athar M, Palle K. Hedgehog/GLI1 Transcriptionally Regulates FANCD2 in Ovarian Tumor Cells: Its Inhibition Induces HR-Deficiency and Synergistic Lethality with PARP Inhibition. Neoplasia. 2021 Sep;23(9):1002–1015. doi: 10.1016/j.neo.2021.06.010. Epub 2021 Aug 8. PMID: 34380074; PMCID: PMC8361230.

37. Mantere T, Tervasmäki A, Nurmi A, Rapakko K, Kauppila S, Tang J, Schleutker J, Kallioniemi A, Hartikainen JM, Mannermaa A, Nieminen P, Hanhisalo R, Lehto S, Suvanto M, Grip M, Jukkola-Vuorinen A, Tengström M, Auvinen P, Kvist A, Borg Å, Blomqvist C, Aittomäki K, Greenberg RA, Winqvist R, Nevanlinna H, Pylkäs K. Case-control analysis of truncating mutations in DNA damage response genes connects TEX15 and FANCD2 with hereditary breast cancer susceptibility. Sci Rep. 2017 Apr 6;7(1):681. doi: 10.1038/s41598-017-00766-9. PMID: 28386063; PMCID: PMC5429682.

38. Meetei AR, de Winter JP, Medhurst AL, Wallisch M, Waisfisz Q, van de Vrugt HJ, … & Wang, W. (2003). A novel ubiquitin ligase is deficient in Fanconi anemia. *Nature Genetics*, 35(2), 165–170. doi: 10.1038/ng1241.

39. Miao, H., Ren, Q., Li, H., Zeng, M., Chen, D., Xu, C., Chen, Y., & Wen, Z. (2022). Comprehensive analysis of the autophagy-dependent ferroptosis-related gene FANCD2 in lung adenocarcinoma. BMC cancer, 22(1), 225. 10.1186/s12885-022-09314-9

40. Moes-Sosnowska J, Rzepecka IK, Chodzynska J, Dansonka-Mieszkowska A, Szafron LM, Balabas A, Lotocka R, Sobiczewski P, Kupryjanczyk J. Clinical importance of FANCD2, BRIP1, BRCA1, BRCA2 and FANCF expression in ovarian carcinomas. Cancer Biol Ther. 2019;20(6):843–854. doi: 10.1080/15384047.2019.1579955. Epub 2019 Mar 1. PMID: 30822218; PMCID: PMC6606037.

41. National Center for Biotechnology Information. ClinVar; [VCV000218824.22], https://www.ncbi.nlm.nih.gov/clinvar/variation/VCV000218824.22 (accessed Oct. 25, 2022).

42. Niraj, J., Färkkilä, A., & D’Andrea, A. D. (2019). The Fanconi Anemia Pathway in Cancer. Annual review of cancer biology, 3, 457–478. 10.1146/annurev-cancerbio-030617-050422

43. Osman, A. E. G., & Deininger, M. W. (2021). Chronic Myeloid Leukemia: Modern therapies, current challenges, and future directions. Blood Reviews, 49, 100825. doi: 10.1016/j.blre.2021.100825.

44. Quintás-Cardama, A., & Cortes, J. (2009). Molecular biology of bcr-abl1-positive chronic myeloid leukemia. Blood, 113(8), 1619–1630. doi: 10.1182/blood-2008-03-144790.

45. R Core Team. (2012). R: A language and environment for statistical computing. R Foundation for Statistical Computing, Vienna, Austria. ISBN 3-900051-07-0, URL http://www.R-project.org/.

46. Senapati, J., Jabbour, E., Kantarjian, H., & Short, N. J. (2023). Pathogenesis and management of accelerated and blast phases of chronic myeloid leukemia. Leukemia, 37(1), 5– 17.

47. Senapati, J., Sasaki, K., Issa, G. C., Lipton, J. H., Radich, J. P., Jabbour, E., & Kantarjian, H. M. (2023). Management of chronic myeloid leukemia in 2023 - common ground and common sense. Blood cancer journal, 13(1), 58.

48. Narlı Özdemir, Z., Kılıçaslan, N. A., Yılmaz, M., & Eşkazan, A. E. (2023). Guidelines for the treatment of chronic myeloid leukemia from the NCCN and ELN: differences and similarities. International journal of hematology, 117(1), 3–15.

49. Eden, R. E., & Coviello, J. M. (2023). Chronic Myelogenous Leukemia. In StatPearls. StatPearls Publishing.

50. Iurlo, A., Cattaneo, D., Consonni, D., Castagnetti, F., Miggiano, M. C., Binotto, G., Bonifacio, M., Rege-Cambrin, G., Tiribelli, M., Lunghi, F., Gozzini, A., Pregno, P., Abruzzese, E., Capodanno, I., Bucelli, C., Pizzuti, M., Artuso, S., Iezza, M., Scalzulli, E., La Barba, G., … Breccia, M. (2023). Treatment discontinuation following low-dose TKIs in 248 chronic myeloid leukemia patients: Updated results from a campus CML real-life study. Frontiers in pharmacology, 14, 1154377.

51. Boucher, L., Sorel, N., Desterke, C., Chollet, M., Rozalska, L., Gallego Hernanz, M. P., Cayssials, E., Raimbault, A., Bennaceur-Griscelli, A., Turhan, A. G., & Chomel, J. C. (2023). Deciphering Potential Molecular Signatures to Differentiate Acute Myeloid Leukemia (AML) with BCR::ABL1 from Chronic Myeloid Leukemia (CML) in Blast Crisis. International journal of molecular sciences, 24(20), 15441.

52. Busch, C., Mulholland, T., Zagnoni, M., Dalby, M., Berry, C., & Wheadon, H. (2023). Overcoming BCR::ABL1 dependent and independent survival mechanisms in chronic myeloid leukaemia using a multi-kinase targeting approach. Cell communication and signaling : CCS, 21(1), 342.

53. Takahashi N. (2023). [Rinsho ketsueki] The Japanese journal of clinical hematology, 64(9), 981–987.

54. Shen Y, Zhang J, Yu H, Fei P. Advances in the understanding of Fanconi Anemia Complementation Group D2 Protein (FANCD2) in human cancer. Cancer Cell Microenviron. 2015;2(4):e986. doi: 10.14800/ccm.986. Epub 2015 Sep 7. PMID: 26640811; PMCID: PMC4667986.

55. Siegel, R.L., Miller, K.D., & Jemal, A. (2017). Cancer Statistics, 2017. CA Cancer J Clin, 67(1), 7–30. doi: 10.3322/caac.21387. Epub 2017 Jan 5. PMID: 28055103.

56. Smogorzewska A, Desetty R, Saito TT, Schlabach M, Lach FP, Sowa ME, Clark AB, Kunkel TA, Harper JW, Colaiácovo MP, Elledge SJ. A genetic screen identifies FAN1, a Fanconi anemia-associated nuclease necessary for DNA interstrand crosslink repair. Mol Cell. 2010 Jul 9;39(1):36–47. doi: 10.1016/j.molcel.2010.06.023. PMID: 20603073; PMCID: PMC2919743.

57. Sokal, J. E., Cox, E. B., Baccarani, M., Tura, S., Gomez, G. A., Robertson, J. E., … & Cervantes, F. (1984). Prognostic discrimination in “good-risk” chronic granulocytic leukemia. *Blood*, 63(4), 789–799.

58. Thompson, L. H., Hinz, J. M., Yamada, N. A., & Jones, N. J. (2005). How Fanconi anemia proteins promote the four Rs: replication, recombination, repair, and recovery. *Environmental and Molecular Mutagenesis*, 45(2-3), 128–142. doi: 10.1002/em.20109.

59. Tsiatis, A. C., Norris-Kirby, A., Rich, R. G., Hafez, M. J., Gocke, C. D., Eshleman, J. R., … & Murphy, K. M. (2010). Comparison of Sanger sequencing, pyrosequencing, and melting curve analysis for the detection of KRAS mutations: diagnostic and clinical implications. *Journal of Molecular Diagnostics*, 12(4), 425–432.

60. Valeri A, Río P, Agirre X, Prosper F, Bueren JA. (2012). Unraveling the role of FANCD2 in chronic myeloid leukemia. *Leukemia*, 26(6), 1447–1448. doi: 10.1038/leu.2012.32.

61. van Twest S, Murphy VJ, Hodson C, Tan W, Swuec P, O’Rourke JJ, … & Deans AJ. (2017). Mechanism of Ubiquitination and Deubiquitination in the Fanconi Anemia Pathway. *Molecular Cell*, 65(2), 247–259. doi: 10.1016/j.molcel.2016.11.005.

62. Venkitaraman, A. R. (2004). Tracing the network connecting BRCA and Fanconi anaemia proteins. Nature Reviews Cancer, 4(4), 266–276. doi: 10.1038/nrc1321.

63. Wang AT, Smogorzewska A. (2015). SnapShot: Fanconi anemia and associated proteins. *Cell*, 160(1-2), 354–354.e1. doi: 10.1016/j.cell.2014.12.031.

64. Wang X, Andreassen PR, D’Andrea AD. (2004). Functional interaction of monoubiquitinated FANCD2 and BRCA2/FANCD1 in chromatin. *Molecular Cell Biology*, 24(13), 5850–5862. doi: 10.1128/MCB.24.13.5850-5862.2004.

65. Williams SA, Wilson JB, Clark AP, Mitson-Salazar A, Tomashevski A, Ananth S, Glazer PM, Semmes OJ, Bale AE, Jones NJ, Kupfer GM. Functional and physical interaction between the mismatch repair and FA-BRCA pathways. Hum Mol Genet. 2011 Nov 15;20(22):4395–410. doi: 10.1093/hmg/ddr366. Epub 2011 Aug 24. PMID: 21865299; PMCID: PMC3196888.

66. World Medical Association. (2007). Declaration of Helsinki 2007. Available online: www.wma.net/e/ethicsunit/helsinki.htm (accessed on 11 February 2021).

67. Xu S, Zhao F, Liang Z, Feng H, Bao Y, Xu W, Zhao C, Qin G. Expression of FANCD2 is associated with prognosis in patients with nasopharyngeal carcinoma. Int J Clin Exp Pathol. 2019 Sep 1;12(9):3465–3473. PMID: 31934192; PMCID: PMC6949855.

68. Xu, J., Wu, M., Sun, Y., Zhao, H., Wang, Y., & Gao, J. (2020). Identifying Dysregulated lncRNA-Associated ceRNA Network Biomarkers in CML Based on Dynamical Network Biomarkers. *BioMed Research International*, 2020, 5189549.

69. Yoshimaru, R., & Minami, Y. (2023). Genetic Landscape of Chronic Myeloid Leukemia and a Novel Targeted Drug for Overcoming Resistance. International journal of molecular sciences, 24(18), 13806.

70. Shin, J. E., Kim, S. H., Kong, M., Kim, H. R., Yoon, S., Kee, K. M., Kim, J. A., Kim, D. H., Park, S. Y., Park, J. H., Kim, H., No, K. T., Lee, H. W., Gee, H. Y., Hong, S., Guan, K. L., Roe, J. S., Lee, H., Kim, D. W., & Park, H. W. (2023). Targeting FLT3-TAZ signaling to suppress drug resistance in blast phase chronic myeloid leukemia. Molecular cancer, 22(1), 177.

71. Li, Z. Y., Zhao, H. F., Zhang, Y. L., & Song, Y. P. (2023). Zhongguo shi yan xue ye xue za zhi, 31(3), 649–653.

72. Telliam, G., Desterke, C., Imeri, J., M’kacher, R., Oudrhiri, N., Balducci, E., Fontaine-Arnoux, M., Acloque, H., Bennaceur-Griscelli, A., & Turhan, A. G. (2023). Modeling Global Genomic Instability in Chronic Myeloid Leukemia (CML) Using Patient-Derived Induced Pluripotent Stem Cells (iPSCs). Cancers, 15(9), 2594.

73. Leung, W., Baxley, R. M., Traband, E., Chang, Y. C., Rogers, C. B., Wang, L., Durrett, W., Bromley, K. S., Fiedorowicz, L., Thakar, T., Tella, A., Sobeck, A., Hendrickson, E. A., Moldovan, G. L., Shima, N., & Bielinsky, A. K. (2023). FANCD2-dependent mitotic DNA synthesis relies on PCNA K164 ubiquitination. Cell reports, 42(12), 113523. Advance online publication.

74. Yilmaz, U., & Eskazan, A. E. (2020). Moving on from 2013 to 2020 European LeukemiaNet recommendations for treating chronic myeloid leukemia: what has changed over the 7 years?. Expert review of hematology, 13(10), 1035–1038.

75. Valeri, A., Alonso-Ferrero, M. E., Río, P., Pujol, M. R., Casado, J. A., Pérez, L., Jacome, A., Agirre, X., Calasanz, M. J., Hanenberg, H., Surrallés, J., Prosper, F., Albella, B., & Bueren, J. A. (2010). Bcr/Abl interferes with the Fanconi anemia/BRCA pathway: implications in the chromosomal instability of chronic myeloid leukemia cells. PloS one, 5(12), e15525.

